# Technical Feasibility Study for a Retinal Camera System

**DOI:** 10.1101/2024.12.20.24319469

**Authors:** Katherine Makedonsky, Anil Patwardhan, Angela Kim, Matthew Silvestrini, Clarissa Lui, Sam Kavusi

**Author notes:** Corresponding author: **Clarissa Lui,** Verily Life Sciences, 999 Bayhill Dre, San Bruno, CA 94066, P., E.

## Abstract

**Background/Objectives:** Improvements in diabetic retinopathy (DR) screening could boost early detection rates and contribute to better patient outcomes. The availability of primary care-based DR screening could address this issue, although operational challenges remain. This study represents a preliminary evaluation of the feasibility and operational performance of a retinal screening system suitable for integration into a primary care setting.

**Methods:** This was a multi-center, data collection study for a non-mydriatic retinal camera system, conducted in 4 iterative development phases; phase 4 simulated a clinical workflow. Study endpoints evaluated image quality (gradability) and operational characterization in phase 4 (time to complete image capture). Participants were required to be at least 22 years of age, and separate participant pools were recruited throughout study phases. In each phase, multiple images per eye were collected, and graded independently by sponsor-employed and external graders.

**Results:** The study included 212 participants. In phases 1-3 (n=192), evaluation of the first image captured per eye for all participants (k=383) by non-sponsor graders showed 93.7% of images (359/383) as gradable; in phase 4 (n=20; k=55), there were 81.8% (45/55) gradable images. Regarding operational results, 90.1% of first-attempts trigger times were less than 30 seconds; by the third attempt, 96.7% were less than 30 seconds,

**Conclusions:** These are encouraging preliminary results in terms of performance and usability for the retinal camera system in this study, which may have potential application in primary care clinics.

## Introduction

Diabetic retinopathy (DR) is a common complication of diabetes mellitus, present in an estimated 25% of all people with diabetes (CDC 2024; Lundeen 2023). DR is a progressive condition that affects the microvasculature of the eye and may lead to irreversible retina damage and ultimately, blindness (Duh 2017). Early DR detection enables timely interventions and can mitigate vision loss in a substantial portion of affected patients, thus the importance of robust screening practices in the population of patients with a history of diabetes (Garg 2009).

However, achieving widespread screening in a large target population poses challenges. In the US, patients may need referrals to ophthalmology clinics, a requirement that can represent a logistical or financial barrier. In fact, the burden associated with specialty care represents a major reason for non-compliance with the American Diabetes Association (ADA) screening guidelines. In underserved areas, where access to properly-equipped ophthalmology clinics is limited, screening rates may drop as low as 20% (Chou 2014; Hartnett 2005; ; Hudson 2022; Chan 2022).

Availability of DR screening in primary care clinics could mitigate this problem. But reports indicate that the success of primary care DR screening programs is uneven and may depend on the size and underlying resource availability of a health organization (Daskivich 2017; Scanlon 2017; Jalkiewicz 2022). Adoption of retinal camera capabilities by primary care clinics requires resource investments and workflow reorganizations. Direct acquisition costs, and requirements to dedicate space and staff represent obstacles to wide deployment of DR screening systems in primary care (Liu 2018; Liu 2019).

These issues provide impetus to design DR screening systems specifically for primary care clinics. These systems would aim to be affordable, easily integrated into primary care workflows and user-friendly for non-dedicated staff, while meeting existing quality standards to produce usable images that are clinically interpretable by ophthalmologists.

We report the results of a prospective study aimed to characterize the performance of a retinal screening camera system for use in a primary care practice. This system, at the time of the study named Verily Retinal Camera (VRC), provides non-mydriatic, color posterior chamber images of the eye that could be suitable for clinical use.

## Methods

### Study Design

This was a multi-center, data collection study consisting of 4 separate phases, which allowed for an iterative approach of device development throughout those phases. The overarching goal was to evaluate the quality of the images captured, and the usability and workflow feasibility of the VRC. The focus areas throughout the phases of the study were image quality, gradability, and functionality of quality control features, as well as operational characterization, i.e., quantification of average attempts and timing needed to obtain acceptable images and complete imaging sessions (end to end).

### Devices

The VRC is a non-mydriatic, 45-degree field imaging retinal camera system that utilizes a set of proprietary machine learning algorithms to generate high fidelity composite color posterior chamber images of the eye by capturing and compiling series of retinal images from each perceived single flash (<200ms). The VRC, consists of a retina camera connected to a laptop computer, where an operator can control both camera and software via the laptop interfaces. Image capture requires a user’s face to be placed on a face rest to look through an optical tube hosting a series of lenses and optical components (beam splitters, mirrors, polarizers, filters, shutters, apertures, motor). Users themselves can focus the image using a knob, before proceeding to align (relying on a gamified user interface that guides head movements until reaching proper position) and center their eye to the optical tube for image capture. During imaging, the retina is illuminated by several LEDs while images are simultaneously captured by an image sensor.

This study investigated several device iterations with different configurations intended to ultimately optimize image gradability. For the majority of the capture session, the illumination is in near infrared range (>850nm), but during the moment of capture, visible white light is used. An OLED micro-display is used to project an image to the retina while stereo pupil tracking cameras track eye position for feedback and alignment guidance to users. The image data is sent to the processor where it is processed into a single retina image.

This study also used reference cameras common in standard ophthalmology clinics, the Nikon Retinastation (RetinaStation 2024), and the CentervueDRSplus (DRSplus 2024) for some of our analyses.

### Participants

Key eligibility criteria were to be at least 22 years of age and provide informed consent, and to not present light hypersensitivity or any contraindication for fundus imaging. While these were the overall criteria, the cohorts enrolled in each one of the study phases were drawn from different population pools (Table S1).

- Phase 1: internal sponsor employees.
- Phase 2: external volunteer participants recruited to the sponsor-base study site (Verily Life Sciences, South San Francisco, CA).
- Phase 3: external volunteer participants recruited to a study site external to the sponsor (Diablo Clinical Research, Walnut Creek, CA).
- Phase 4: external volunteer participants recruited to the study site external to the sponsor (as in phase 3). The differences with phase 3 were in the camera system (which had a quality control feature incorporated) and in the study procedures (which were conducted under a simulated clinical environment).

The study received Institutional Review Board (IRB) approval from Western IRB prior to initiation. All participants signed informed consent approved by the IRB.

### Procedures

#### Study visits

For Phase 1 of the study, participants provided demographic information, but not self-reported medical history. We collected up to 6 images using two camera configurations. Then, participants took a survey questionnaire about their experience. Finally, operators captured images with the DRSPlus system, up to 3 per eye, at their discretion (without further recapture after obtaining an image of appropriate quality).

For Phase 2, study procedures were similar to Phase 1. Thus, the main difference between Phases 1 and 2 was the type of participant recruited, which enabled collection of self-reported medical history information (from non-employees) during Phase 2.

For Phase 3, study procedures included collection of self-reported medical history information. Images from the VRC were collected only with one camera configuration, 3 times per eye, in order to maximize the number of images collected. Images were also collected from two reference systems, the Centervue DRSplus and the Nikon Retinastation, up to 3 per eye. During Phase 4, the underlying goal of the study procedures was to simulate a clinical workflow. This phase introduced a VRC feature called the “image quality control feature (IQCF)” which scored quality in real time during capture (giving a cue of whether recapture was needed or not). Participants signed consent, and provided demographics and self-reported medical history. Operators captured up to 3 images with VRC, at their discretion (as it would be in an actual clinical setting) and guided by the quality scores generated by the IQCF on whether recaptures were needed. The timing of the session was recorded from start to finish. After completing the user survey, reference images were captured with the Retinastation system.

#### Grading

For Phases 1-2, an sponsor-employed optometrist evaluated gradability of all VRC images based on guidelines set forth by an independent third-party teleretinal grading organization :

- Focus/blur: gradable when the third order branch is visible and clear.
- Optic Disc: gradable when the optic disc is 100% visible.
- Fovea: gradable when at least 1 disc size around the fovea is present.
- Field of view: gradable when at least two thirds of the field of view are visible.

In addition, in Phases 1-3, the first VRC image per eye per participant was evaluated for gradability by non-sponsor graders. Three independent graders from a third-party independent teleretinal grading organization evaluated images; final gradability per image was determined by majority vote of graders.

For phase 4, as part of the clinical workflow simulation, all VRC acquired images were sent to an independent third-party teleretinal grader, but they were evaluated by only one individual grader in this case.

The sponsor-employed optometrist evaluated the images from the reference devices across all study phases; those images were not sent for third-party teleretinal grading.

### Analyses

An original sample size estimate of at least 125 was proposed for the purpose of image quality evaluation. Considering other objectives in addition to image quality (ie, simulating a clinical workflow to assess operational aspects), the study enrolled a total of 212 participants to aim for a diverse cohort base, with both diseased and healthy eyes. Overall, this was an iterative early feasibility study, therefore sample sizes for each phase varied based on their specific aims.

The key gradability-based endpoint was the percentage of individual images classified as gradable by the corresponding graders in each phase (sponsor employee in phases 1-2, external graders in phases 1-4). In addition, we evaluated the percentage of gradable images according to factors expected to affect gradability, such as pupil size or presence of cataracts.

The key operations-based endpoint was time, divided in segments. “Trigger time” was the time to achieve optimal alignment in each attempt before triggering the flash, for each participant; for each attempt, time was averaged between right and left eye. “Complete workflow time” was the time lapsed from the time the participant sat in front of the device until the final image capture was completed, we analyzed this endpoint in Phase 4 only (the one conducted under simulated clinical workflow conditions).

Overall, we present pooled results for phases 1-3 (which were conducted in largely comparable environments), and separately for phase 4 (which was conducted differently, with the IQCF incorporated in the camera system, and overall in a simulated clinical setting).

## Results

### Characteristics of Study Cohorts

The study was conducted between April and September of 2022. There were a total of 212 participants enrolled, 192 pooled into Phases 1-3, and 20 for Phase 4 (Table 1). Of the patients with a self-reported history of diabetes, most had had it for longer than 15 years.

**Table 1.** Demographic and relevant medical characteristics of the study cohorts.

|  |  | Phase 1-3 (n=192) | Phase 4 (n=20) |
| --- | --- | --- | --- |
| Age, yrs | Mean (SD) | 54.80 (16.57) | 48.95 (10.30) |
|  | Min-Max | 23.00-91.00 | 33.00-61.0 |
| Sex, n (%) | Female | 94 (49.0) | 11 (55.0%) |
|  | Male | 98 (51.0) | 9 (45.0%) |
| Gender, n (%) | Man | 98 (51.0) | 9 (45.0%) |
|  | Woman | 94 (49.0) | 11 (55.0%) |
| Ethnicity, n (%) | Not of Hispanic, Latino/a, or Spanish Origin | 168 (87.5) | 20 (100.0%) |
|  | Of Hispanic, Latino/a, or Spanish Origin | 24 (12.5) | 0 |
| Race, n (%) | Asian | 44 (22.9) | 1 ( 5.0) |
|  | Black African American | 18 ( 9.4) | 1 ( 5.0) |
|  | Other | 2 (1) |  |
|  | White | 115 (59.9) | 18 (90.0) |
|  | Decline To Answer | 6 ( 3.1) |  |
|  | Multiple Race | 7 ( 3.6) |  |
| Diagnosed diabetes, n (%) | Yes | 123 (64.1) | 20 (100) |
|  | No | 23 (12) | 0 |
|  | NA | 46 (24) | - |
| Diabetes diagnosis duration | Mean, yrs | 21.3 (13.0) | 25.7 (12.4) |
|  | Q1-Q3, yrs | - | 15.3-33.3 |
|  | Min-Max, yrs | 1-64.5 | 6.0-47.0 |
|  | <5 yrs, n (%) | 8 ( 4.2) | 0 |
|  | 5-10 yrs, n (%) | 10 ( 5.2) | 3 (15.0) |
|  | 10-15 yrs, n (%) | 17 ( 8.9) | 2 (10.0) |
|  | 15+ yrs, n (%) | 88 (45.8) | 15 (75.0) |
|  | NA, n (%) | 69 (35.9) | - |
| Diagnosed<br>cataracts | Don 't know | 90 (46.9) | 20 (100) |
|  | No | 33 (17.2) | 0 |
|  | Yes | 23 (12.0) | 0 |
|  | NA | 46 (24.0) | - |
| Cataract<br>surgery | Don 't know | 90 (46.9) | 20 (100) |
|  | No | 49 (25.5) |  |
|  | Yes | 7 ( 3.6) |  |
|  | NA | 46 (24.0) |  |

### Image Quality and Gradability

While we collected VRC images using two configuration setups during study phases 1-2 (with k=209 and 475, respectively), we found no meaningful difference in gradability (evaluated by a sponsor-employed grader) between the two configurations. More than 75% of images for both configurations were deemed gradable, with 6%-8% of images considered as ‘borderline.’ Based on these findings, we focused on using only one configuration for the subsequent phases 3-4.

Evaluation of the first image captured per eye for all participants (N=192; k=383) in phases 1-3 by non-sponsor graders showed 93.7% of images (359/383) as gradable and 6.3% (24/383) as ungradable (Table 2).

**Table 2.** Summary of gradability results.

|  |  | Phases 1-3 (k=383) | Phase 4 (k=55 ) |
| --- | --- | --- | --- |
| Gradable, k (%) |  | 359 (93.7) | 45 (81.8) |
| Ungradable, k (%) |  | 24 (6.3) | 10 (18.2) |
| Gradable by pupil size, k (%) | < 2.5 mm | 0/11 | 3/6 (50.0) |
|  | [2.5, 3.0] mm | 6/8 (75.0) | - |
|  | [3.0, 3.5] mm | 15/21 (71.4) | - |
|  | [3.5, 4.0] mm | 30/32 (93.8) | 1/4 (25.0) |
|  | [4.0, 4.5] mm | 28/28 (100.0) | 6/9 (66.7) |
|  | [4.5, 5.0] mm | 67/67 (100.0) | 9/9 (100.0) |
|  | >= 5.0 mm | 213/216 (98.6) | 26/27 (96.3) |
| Gradable in participants with history of cataracts, k (%) |  | 39/45 (86.7) | – |

We also collected images from reference systems in order to have contextual information for our procedures and our observations. The sponsor-employed grader evaluated the first image generated by reference camera systems from the phase 3 of the study (n=107; k=214). We focused on this phase, since it was the one where 2 reference systems were deployed and where all participants self-reported a diabetes diagnosis. The Nikon Retinastation device had 67% gradable images (144/214). The operator could not acquire images for all participants, missing 18% of images (which were computed as ‘ungradable’), from participants with small pupils. The DRSplus device had 84% gradable images (184/214 total images).

To better understand the impact of specific factors on gradability, we analyzed the percentage of gradable images across phases 1-3 according to pupil size and cataract history (Table 2). Graders labeled as gradable 52.5% (21/40) of images obtained with pupils<3.5 mm, and 98.5% (338/343) of those from pupil ≥3.5mm. In addition, images from participants with cataract history had overall 86.7% gradability (39/45).

We then assessed the percentage of gradable images captured in conditions simulating a clinical workflow (phase 4 of the study; n=20; k=55), where all images acquired per participant were sent to one grader. There were 81.8% (45/55) gradable images and 18.2% (10/55) ungradable images. Evaluated on a participant level (since multiple images were sent per participant), 85% (17/20) of participants had gradable images for both eyes. Consistent with the results from phase 1-3, the percentage of gradable images was lower with pupils <3.5 mm (4/10, 40%).

Graders were able to render diagnoses for 196 participants (used only for research purposes). Of the 143 participants diagnosed with diabetes, 40 participants (28%) had retinopathy (Table S2).

### System Operation

Each participant in phases 1-3 got three VRC image captures per eye. Trigger times decreased with each attempt, and, by attempt 3, there was less variation between participants. On the first attempt 90.1% of participants had a trigger time less than 30 seconds, and by the third attempt, 96.7% of participants had a trigger time less than 30 seconds (Table 3). The age of participants affected their trigger time: first-attempt trigger times were <10 seconds for a majority of participants under 30 years (63.2%, 12/19), but for a lower proportion of participants over 65 years (29%, 18/62).

**Table 3.**
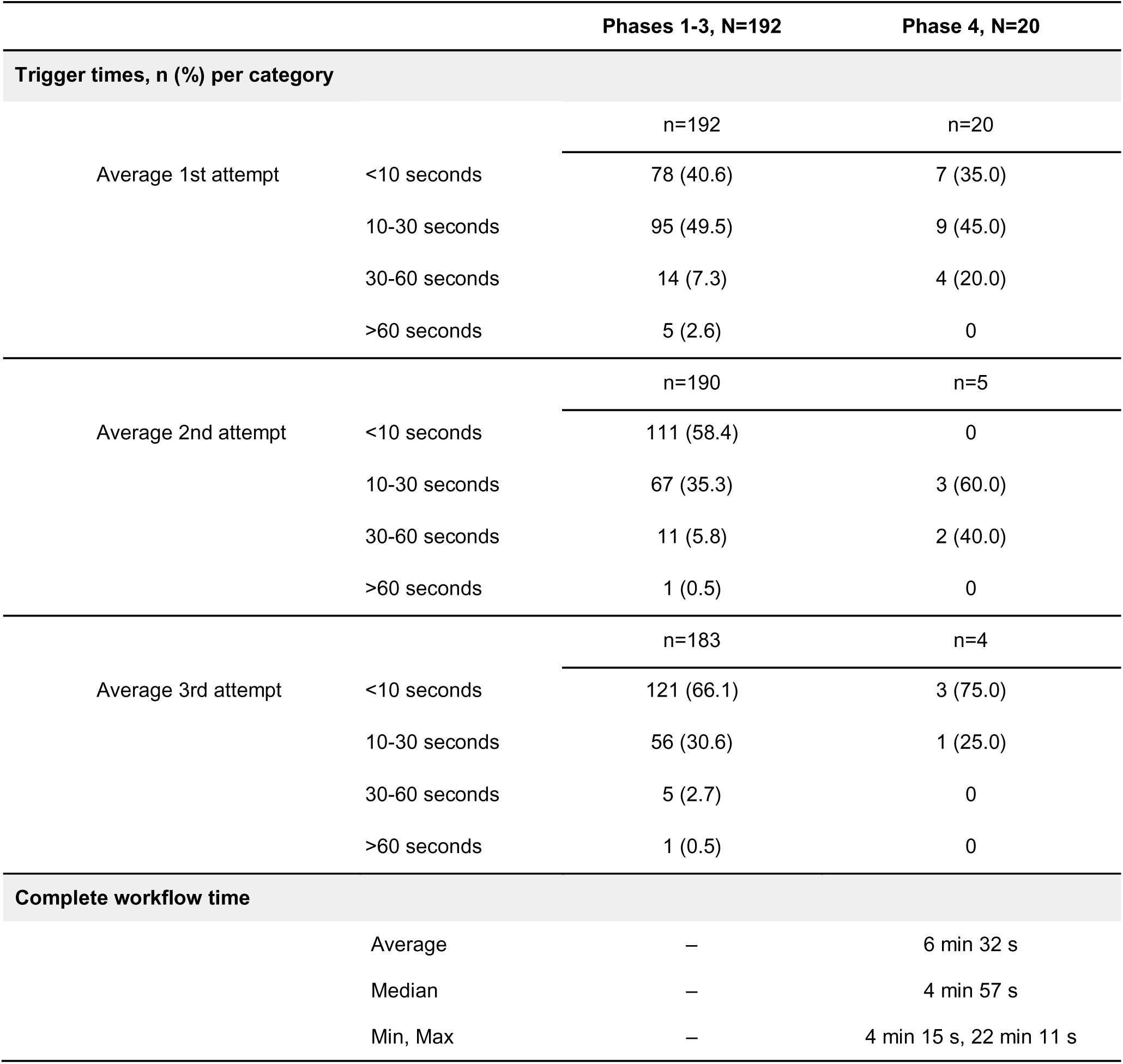
Summary of operation time results across study phases.

Not all participants in phase 4 took three attempts to capture images. The overall results regarding trigger times appear to show trends (as cohort sizes were small) consistent with the observations from phases 1-3, in that attempts progressively shortened for most participants. The median duration of a complete session was 4 minutes 57 seconds.

## Discussion

We report herein results from a feasibility study showing that the VRC can produce retinal images of quality enough to be gradable in clinical practice, overall more than 90% of the VRC images were classified as gradable by human graders. Procedures to operate the VRC were accessible for most users, as more than 90% of participants could capture images in less than 30 seconds in their first attempt.

In this study, we set out to investigate two critical features for a retinal camera system that aspires to be effectively integrated into primary care settings. One aspect is ensuring that the performance of the device will be appropriate and the images produced could actually be utilizable in the clinic. In that regard, this device showed promising results (particularly for a non-mydriatic system) appearing capable to meet expected standards of performance quality for retinal cameras, producing between 80%-90% of gradable images (Fenner 2018; Piyasena 2019; Davila 2017; RCOphth 2012; Rajalakshmi 2021; Scanlon 2017). We did perform analyses in parallel with other camera systems already available in clinics. While these were not formal comparative analyses, our findings are encouraging showing that the proportions of gradable images from the VRC may be numerically similar to these standard systems.

The second aspect of interest is to maximize ease of operations by non-dedicated staff, as hypothetical operators in primary care clinics would be. We showed that the duration of imaging sessions with untrained or lightly-trained operators was within what could be considered reasonable for the intended setting. There was a noticeable learning effect in real time as operators/users repeated attempts at increasingly faster pace. This further supports the notion that this can be a manageable and low-burden system, feasible to be used by untrained or lightly trained personnel that can refine their skills quickly, and suitable to be integrated in primary care settings.

Our results are promising, but we acknowledge that this study has limitations. Our study design was exploratory in nature, so it does not represent a formal validation of the VRC. We followed a design that allowed us to test multiple configuration setups in iterative fashion; we could make decisions about pursuing or discarding certain design elements, but this approach precluded a deeper characterization of particular system setups at scale enough to extract robust conclusions. We collected limited information from study participants, in terms of relevant medical history and clinical characteristics; therefore, we cannot determine to which extent, if at all, our participants were representative of the target population of patients with diabetes in primary care clinics. It remains to be determined whether performance will be consistent across cohorts of individuals diverse in terms of visual acuity, severity of diabetes diagnosis, or presence of other ophthalmologic issues. Altogether, these limitations point to future studies necessary to fully characterize the VRC, such as formal and appropriately powered comparisons with state-of-art equipment, investigations set up to understand performance across diverse environments, and with more diverse users and operators.

In summary, this report presents encouraging preliminary results in terms of performance and usability for the VRC, a retinal camera system with potential application in primary care clinics. Our findings justify subsequent studies to fully characterize its performance. This device may represent an advance towards wider availability of retinal screening in primary care settings, a welcome development with the potential to mitigate morbidity and improve outcomes for patients with diabetes.

## Prior disclosure of these data

Not applicable

## Data sharing statement

The data supporting this study are not available for sharing

## Authors’ contributions

Study concept and design: KM, AP, CL, SK

Data collection: AK

Data analysis and interpretation: AP, KM

Draft writing and review: All

Draft approval for submission: All

## Acknowledgements

Authors wish to acknowledge writing and editing support from Julia Saiz from Verily Life Sciences.

## SUPPLEMENT/APPENDIX

**Table S1.** Summary of study elements and procedures, according to phase.

|  | Phase 1 | Phase 2 | Phase 3 | Phase 4 |
| --- | --- | --- | --- | --- |
| Participants | Sponsor employees<br>n=46 | External<br>n=39 | External<br>n=107 | External<br>n=20 |
| Collected demographic info. | Yes | Yes | Yes | Yes |
| Collected self-reported medical history info. | No | Yes | Yes | Yes |
| Study site | Sponsor location | Sponsor location | External site | External site<br>Simulated clinic environment |
| Number of VRC images taken | 3/eye with each configuration | 3/eye | 3/eye | Up to 3/eye<br>(guided by IQCF) |
| Reference system images taken | DRSPlus<br>Up to 3/eye | DRSPlus<br>Up to 3/eye | DRSplus<br>Nikon Retinastat.<br>Up to 3/eye. | Nikon Retinastat.<br>Up to 3/eye |
| Sponsor employee grader | Yes | Yes | Yes | Yes |
| Images graded | All, VRC and ref | All, VRC and ref | All, VRC and ref | All, VRC and ref |
| External graders | Yes<br>Committee of 3 | Yes<br>Committee of 3 | Yes<br>Committee of 3 | Yes<br>Single grader |
| Images graded | 1st VRC image | 1st VRC image | 1st VRC image | All VRC images |
| Time measured |  |  |  |  |
| Trigger time | Yes | Yes | Yes | Yes |
| Complete time | No | No | No | Yes |

**Table S2.** Diagnoses generated by graders, based on VRC images.

| Reference Diagnosis, n (%) | Total Participants<br>(N=212) |
| --- | --- |
| Ungradable | 16 (7.5) |
| Mild Diabetic Retinopathy | 8 (3.8) |
| Moderate Diabetic Retinopathy | 16 (7.5) |
| Severe Diabetic Retinopathy | 1 (0.5) |
| Proliferative Diabetic Retinopathy | 15 (7.0) |
| Other Conditions (based on retina photos) | 11 (5.2) |

